# Vaccine effectiveness against COVID-19 related hospital admission in the Netherlands: a test-negative case-control study

**DOI:** 10.1101/2021.11.09.21266060

**Authors:** F.A. Niessen, Mirjam J. Knol, Susan J.M. Hahné, VECTOR study group, M.J.M. Bonten, P.C.J.L. Bruijning-Verhagen

**Affiliations:** Julius Center for Health Sciences and Primary Care, University Medical Center Utrecht, Utrecht, Netherlands; Centre for Infectious Disease Control, National Institute for Public Health and the Environment, Bilthoven; Catharina Ziekenhuis, The Netherlands; Medisch Spectrum Twente, The Netherlands; Onze Lieve Vrouwe Gasthuis, The Netherlands; Martini Ziekenhuis, The Netherlands; St. Antonius Ziekenhuis, The Netherlands; Canisius Wilhelmina Ziekenhuis, The Netherlands

## Abstract

**Introduction:** Real-world vaccine effectiveness (VE) estimates are essential to identify potential groups at higher risk of break-through infections and to guide policy. We assessed the VE of COVID-19 vaccination against COVID-19 hospitalization, while adjusting and stratifying for patient characteristics.

**Methods:** We performed a test-negative case-control study in six Dutch hospitals. The study population consisted of adults eligible for COVID-19 vaccination hospitalized between May 1 and June 28 2021 with respiratory symptoms. Cases were defined as patients who tested positive for SARS-CoV-2 by PCR during the first 48 hours of admission or within 14 days prior to hospital admission. Controls were patients tested negative at admission and did not have a positive test during the 2 weeks prior to hospitalization. VE was calculated using multivariable logistic regression, adjusting for calendar week, sex, age, comorbidity and nursing home residency. Subgroup analysis was performed for age, sex and different comorbidities. Secondary endpoints were ICU-admission and mortality.

**Results:** 379 cases and 255 controls were included of whom 157 (18%) were vaccinated prior to admission. Five cases (1%) and 40 controls (16%) were fully vaccinated (VE: 93%; 95% CI: 81 – 98), and 40 cases (11%) and 70 controls (27%) were partially vaccinated (VE: 70%; 95% CI: 50-82). A strongly protective effect of vaccination was found in all comorbidity subgroups. No ICU-admission or mortality were reported among fully vaccinated cases. Of unvaccinated cases, mortality was 10% and 19% was admitted at the ICU

**Conclusion:** COVID-19 vaccination provides a strong protective effect against COVID-19 related hospital admission, in patients with and without comorbidity.

## Introduction

Only about a year after SARS-CoV-2 emerged as a new respiratory virus, the first vaccines were approved, based on licensure trials yielding high vaccine efficacy against symptomatic SARS-CoV-2 infection, especially for mRNA-vaccines (1-3).

In the Netherlands, COVID-19 vaccination started early January 2021. Elderly living in nursing homes, their caregivers and acute care workers were the first groups to receive vaccination. Vaccine roll-out then followed an age-stratified approach, starting with elderly >90 years of age, followed by successively younger 5-year age groups. In addition, people <60 years with an increased risk of severe disease due to obesity, chronic pulmonary disease, chronic cardiac disease, diabetes and immune deficiency were prioritized. As of 12 October 2021, in the Netherlands, 10,8 million citizens aged > 11 years have been fully vaccinated resulting in a vaccination coverage of 82.4% in the adult population (4). Four vaccines are used in the Netherlands: Spikevax©, Comirnaty©, Vaxzevria© and COVID-19 Vaccine Janssen©.

Once vaccination has been implemented, monitoring vaccine effectiveness (VE) is needed to measure the real life effectiveness of vaccination under changing epidemiological conditions (e.g. emerging variants) and for different vaccines, to identify potential groups at higher risk of break-through infections and to guide policy making on additional vaccination and booster vaccination. The test-negative case-control design is well suited for quantifying VE and is therefore often used in post-licensure VE studies because of its efficiency and limited bias compared to other observational designs (5). Importantly, in this design bias due to health seeking behavior or access to health care and vaccination is limited by only including patients seeking care for a specified set of symptoms. Yet, comorbidities can be important confounders, and many observational studies on VE of COVID-19 vaccination lack details on patient comorbidity status as such data cannot be comprehensively collected without detailed chart review. Furthermore, stratified estimates for different risk-groups are essential for further decision making regarding additional COVID-19 vaccination. The aim of this study was to determine the VE of COVID-19 vaccination against COVID-19 hospitalization stratified for comorbidity and with adjustment for relevant patient characteristics.

## Methods

### Design and data collection

A test-negative case-control study was performed in six Dutch hospitals geographically distributed across the Netherlands. The study population consisted of adults admitted to the hospital with respiratory symptoms and eligible for COVID-19 vaccination at the time of admission. Inclusion criteria were cough and/or shortness of breath with an onset within 21 days before hospitalization, and a respiratory specimen tested for SARS-CoV-2 (through PCR/LAMP-PCR) at admission or in the 14 days prior to admission at the municipal health services. Vaccine eligibility was determined based on the age-stratified roll-out of the Dutch COVID-19 immunization program. We distinguished three age-categories with an inclusion period based on vaccine eligibility. Patients born before 1947 and admitted between March 1 and May 10, patients born from 1946 to 1960 admitted between April 12 and June 7 and patients born from 1961 to 2003 admitted between May 3 and July 5 were included. Patients were screened retrospectively by chart review for in- and exclusion criteria. Each respective week the first 3-5 eligible patients in each age category were included per hospital. Patients transferred from another hospital for possible or confirmed COVID-19 and patients readmitted within 14 days of a prior hospitalization were not included. Patients without information on vaccination status and/or on their SARS-CoV-2 test results were excluded from the analysis. Controls without negative test results at hospital admission were also excluded.

Clinical data including demographic characteristics, medical history and medication, COVID-19 test results, COVID-19 vaccination status and disease course were collected by retrospective chart review by dedicated research personnel and recorded in an electronic case report form. Vaccination status, including date of vaccination and vaccine type, was verified and complemented with the National vaccination registry (CIMS), containing data on COVID-19 vaccination of all Dutch citizens who provided consent; it is estimated that 93% of vaccinated persons give informed consent.

### Definitions

Cases were defined as patients hospitalized with cough and/or dyspnea with a respiratory sample positive for SARS-CoV-2 by PCR/LAMP-PCR tested within 48 hours after hospital admission or at the municipal health services no more than 14 days preceding hospital admission. Controls were defined as patients hospitalized with cough and/or dyspnea testing negative for SARS-CoV-2 by PCR/LAMP-PCR on all respiratory samples taken during the first 48 hours of admission and in the 14 days preceding hospital admission.

Individuals were considered fully vaccinated if they had received both doses of a two-dose vaccine (Spikevax©, Comirnaty©, Vaxzevria©) schedule at least 14 days before symptom onset, or one dose of a single-dose vaccine (COVID-19 Vaccine Janssen©) schedule at least 28 days before symptom onset. Participants who received one dose of a two-dose vaccine at least 14 days before symptom onset, were considered partially vaccinated. All patients who did not receive a COVID-19 vaccine or received a first dose less than 14 days prior to symptom onset (or <28 days for vaccination with a single-dose vaccine) were considered unvaccinated. No data was available on prior SARS-CoV-2 infection.

Data on comorbidities were collected through chart review. We classified comorbidities based on underlying conditions as recorded in medical records combined with data on chronic medication. Obesity was defined as a Body Mass Index >= 30. Patients were considered immunocompromised when using immunosuppressant medication (systemic corticosteroids or disease modifying antirheumatic drugs), having end stage renal failure or having an immune deficiency disorder. Patients with recorded chronic pulmonary disease, asthma or using chronic inhalation corticosteroids were considered as chronic respiratory patients.

### Statistical analysis

The primary objective was to estimate VE against COVID-19 related hospital admission for fully and partially vaccinated subjects. Secondary objectives were to estimate VE by age group and comorbidities and to estimate VE against ICU-admission and in-hospital mortality.

Patient characteristics for cases and controls were described and differences were tested using the Mann Whitney U test for continuous variables, Fishers exact test for proportions and chi square test for categorical variables. Because the inclusion-procedure was stratified by age, the overall VE-estimates were calculated by weighing the age stratified estimates respective to the age distribution of COVID-19 hospitalization in the general population.

VE was calculated for both full and partial vaccination as preventive factor for COVID-19 hospitalization by 1 minus the odds ratio (OR). The OR and 95% CI was calculated using logistic regression adjusted for age group, week of symptom onset (cubic spline), sex, nursing home residency and underlying diseases. We took into account diabetes, chronic cardiac disease, chronic pulmonary disease, immune deficiency, malignancy and obesity as underlying diseases, since these were associated with vaccination and severe outcome (6-8). Overall VE as well as product specific VE were estimated. Subgroup analysis were performed for age, sex and comorbidity. To assess whether there was effect modification by age or sex, separate models were fit with interaction terms for these covariables. For ICU admission and mortality, we performed a descriptive analysis and compared baseline characteristics and outcomes in cases by vaccination status.

Several sensitivity analyses were performed on the primary outcome. First, VE was calculated among inclusions meeting the WHO SARI-criteria (acute onset cough and fever) (9). Second, patients with unknown vaccination status were considered as non-vaccinated to check for potential bias due to missing information. Third, we considered vaccination to be protective after 7 days instead of 14 days for 2-dose vaccines and 14 days instead of 28 days for the 1-dose vaccine.

## Results

### Study population

Between March 1 and June 26 2021 a total of 678 patients were included in six hospitals. Of these patients, 44 were excluded for analysis due to missing data on vaccination status (n=38) and/or SARS-CoV-2 status (n=6). The remaining 634 inclusions consisted of 379 patients positively tested on SARS-CoV-2 (cases) and 255 controls. Characteristics of the cases and controls are shown in table 1. The median age was 68 years (IQR 56–78) and was lower in cases than controls (66 vs 71; p<0.05). Since the distribution over the three age groups was comparable to the age-distribution in hospitalized COVID-19 patients in the Netherlands, weighing was considered not to be of additional value (10). Most patients (82%) had at least one comorbidity. Obesity was more frequent among cases than controls (38% vs 23%), whereas other underlying conditions like chronic pulmonary disease, immune deficiency, chronic cardiac disease and malignancy were more frequent among controls (p<0.05). Fever at or preceding hospital admission was reported by 239 cases (63%) and 109 controls (43%). Median CRP was 100 Ug/l among cases and 61 Ug/l among controls. Median leukocyte counts were 6.6 *10^9/L and 12.0 *10^9/L among cases and controls, respectively. Controls were more frequently also tested for other respiratory pathogens (86%, n=218) than cases (30%, n=113). Based upon these tests, seven controls had evidence of infection caused by other respiratory pathogens: *Streptococcus pneumoniae* (n=4), influenza (n=1), respiratory syncytial virus (n=1), and rhinovirus (n=1). One case also tested positive for rhinovirus. Vaccination status for influenza and *Streptococcus pneumoniae* was only documented for 1% of the cases and 3% of the controls.

**Table 1:** patient characteristics

|  | **Controls**<br>**N = 255** | **Cases**<br>**N= 379** |  |
| --- | --- | --- | --- |
| **Median age (IQR)** | 71.00 [61, 83] | 66.00 [56, 76] | <0.001 |
| **Age group** |  |  |  |
| **Age > 75** | 112 (44) | 110 (29) |  |
| **Age 61-75** | 81 (32) | 134 (35) |  |
| **Age 18-60** | 62 (24) | 135 (36) |  |
| **Sex (male)** | 136 (53) | 228 (60) | 0.105 |
| **Long term care facility resident** |  |  | 0.022 |
| **Yes** | 14 (6) | 8 (2) |  |
| **Unknown** | 8 (3) | 5 (1) |  |
| **Median BMI (IQR)** | 26.7 (5.3) | 29.4 (5.7) | <0.001 |
| **Chronic respiratory disease** | 120 (47) | 100 (26) | <0.001 |
| **Chronic cardiac disease** | 88 (35) | 86 (23) | 0.001 |
| **Immunocompromised** | 50 (19.6) | 35 (9.2) | <0.001 |
| **Malignancy** | 47 (18) | 32 (8) | <0.001 |
| **Obesity** | 58 (23) | 143 (38) | <0.001 |
| **Diabetes** | 47 (18) | 108 (29) | 0.005 |
| **Hypertension** | 88 (35) | 126 (33) | 0.807 |
| **Other** | 158 (62) | 214 (57) | 0.195 |
| **Multi comorbidity** | 128 (50) | 146 (39) | 0.005 |
| **No comorbidity** | 22 (9) | 47 (12) | 0.172 |
| **Smoking status known** | 39 (15) | 73 (19) | 0.239 |
| **Current smoker** | 62 (24) | 37 (10) | <0.001 |
| **Ex-smokers** | 99 (39) | 133 (35) | 0.383 |
| **Median days between symptom onset and hospital admission (IQR)** | 2.00 [1, 5] | 7.00 [4, 9] | <0.001 |
| **Fever** | 109 (43) | 239 (63) | <0.001 |
| **Cough** | 168 (66) | 296 (78) | 0.001 |
| **Dyspnea** | 231 (91) | 338 (89) | 0.661 |
| **CRP Median concentration in Ug/l** | 61 [17, 141] | 100 [50, 155] | <0.001 |
| **Leucocytes Median concentration in 10<sup>9</sup>/L** | 12.0 [8.8, 14.8] | 6.63 [4.8, 9.3] | <0.001 |
| **SARS-CoV-2 serology test** | 16 (6) | 92 (24) | <0.001 |
| **Positive SARS-CoV-2 serology** | 5 (31) | 77 (84) | <0.001 |
| **Tested at other respiratory pathogens at admission** | 218 (86) | 113 (30) | <0.001 |
| **Positive for other resp. pathogen** | 7 (3) | 1 (1) | 0.353 |

### Vaccine effectiveness

In total 110 patients (17%) were partially vaccinated (40 cases (11%) and 70 controls (27%)) and 45 were fully vaccinated (5 cases (1%) and 40 controls (16%)) (Table 2). VE was adjusted for week of symptom onset and age group was 66% (95% CI: 46-79) for partial vaccination and 93% (95% CI: 82 – 98) for full vaccination. With additional adjustment for sex, comorbidity and nursing home residency, VE was 70% (95% CI: 50 – 82) for partial vaccination and 93% (95% CI: 80 – 98) for full vaccination (Table 2). Sensitivity analyses showed higher VE-estimates when using a more infection-specific case definition and lower VE-estimates when shortening the interval between disease onset and vaccination (Table 1 in supplement).

**Table 2:** Overall VE and VE by vaccine product adjusted for age, week of symptom onset, nursing home residency and comorbidity

|  |  |  | VE adjusted for age group<br>and week of symptom<br>onset |  | Fully adjusted<br>VE* |  |
| --- | --- | --- | --- | --- | --- | --- |
|  | Cases | Controls | VE % | 95% CI | VE % | 95% CI |
| **Overall** |  |  |  |  |  |  |
| Unvaccinated | 334 | 145 |  |  |  |  |
| Partially vaccinated | 40 | 70 | **66** | (46 - 79) | **70** | (50 - 82) |
| Fully vaccinated | 5 | 40 | **93** | (82 - 98) | **93** | (80 - 98) |
| **Comirnaty** |  |  |  |  |  |  |
| Partially vaccinated | 22 | 44 | **68** | (43 - 82) | **69** | (41 - 84) |
| Fully vaccinated | 4 | 30 | **94** | (81 - 98) | **94** | (80 - 98) |
| **Vaxzevria** |  |  |  |  |  |  |
| Partially vaccinated | 12 | 15 | **65** | (15 - 86) | **70** | (25 - 88) |
| Fully vaccinated | 0 | 1 | **100** |  | **100** |  |
| **Spikevax** |  |  |  |  |  |  |
| Partially vaccinated | 3 | 7 | **75** | (-10 - 94) | **81** | (-1 - 97) |
| Fully vaccinated | 0 | 8 | **100** |  | **100** |  |

\* adjusted for time (week of symptom onset), age, sex, nursing home residency, diabetes, chronic cardiac disease, chronic pulmonary disease, immune deficiency, malignancy and obesity as underlying disease.

Of all vaccinated patients the majority (67%) was vaccinated with Comirnaty, 18% with Vaxzevria, 12% with Spikevax and 1% (n=2) with COVID-19 vaccine Janssen. In seven patients the vaccine-product was unknown. For Comirnaty the product specific VE was 69% and 94% for partial and full vaccination, respectively (Table 2). VE was high for all vaccines and for all age groups with no statistical interaction between age groups and vaccination status, although numbers were low especially in the 18-60 year age group. Eighty percent of fully vaccinated cases were female and VE for both partial and full vaccination tended to be higher in males than in females though this difference was not statistically significant. Subgroup analyses for different comorbidities showed a protective effect of both full and partial vaccination among all comorbidities (figure 1). VE-estimates for full vaccination were above 90% for all comorbidities except among immunocompromised (VE 75%; 95% CI: −177 – 98).

**Figure 1:**
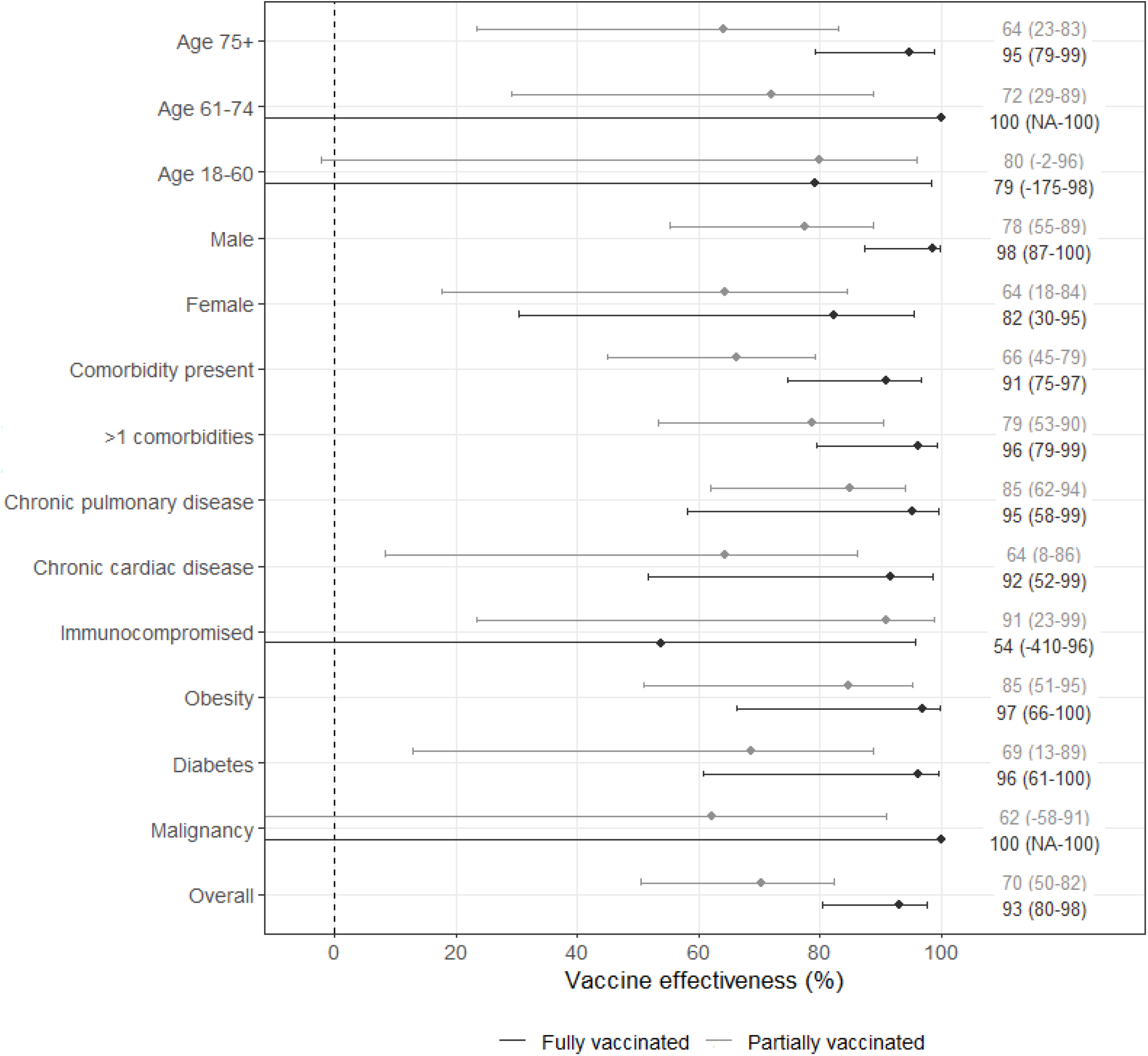
Effectiveness of partial and full vaccination, stratified by age-group, sex and comorbidity, adjusted for week of symptom onset, age, sex, nursing home residency and comorbidity.

### Vaccinated vs unvaccinated cases

Characteristics of vaccinated cases compared to unvaccinated cases are described in table 3.. Among the fully vaccinated cases, time since last vaccination ranged between 17 and 62 days (median of 33 days). Median age was higher in fully vaccinated cases compared to partially and unvaccinated cases. All vaccinated cases had at least one comorbidity, of the unvaccinated cases only 86%had a comorbidity. In addition to female sex (80%), obesity (60%) and immunocompromising conditions (40%) were more frequent among fully vaccinated cases. Median length of stay was the same in vaccinated, partially vaccinated and unvaccinated cases. Of all cases, 69 were admitted in ICU (18%), 64 (19%) of unvaccinated cases and 5 (13%) of partially vaccinated cases. Forty-two cases died during hospitalization, 33 (10%) of the unvaccinated cases and 9 (23%) of the partially vaccinated cases. No ICU-admission or in-hospital mortality was reported among fully vaccinated cases.

**Table 3:** Characteristics of unvaccinated, partially vaccinated and fully vaccinated cases

|  | Fully vaccinated cases | Partially vaccinated cases | Unvaccinated cases |
| --- | --- | --- | --- |
| **n** | 5 | 40 | 334 |
| **Age median (IQR)** | 85 [60, 86] | 78 [62, 84] | 65 [55, 75 ] |
| **Age group** |  |  |  |
| **75+** | 3 (60) | 20 (50) | 87 (26) |
| **61-74** | 0 (0) | 16 (40) | 118 (35) |
| **18-60** | 2 (40) | 4 (10) | 129 (37) |
| **Male Sex** | 1 (20) | 25 (63) | 202 (61) |
| **Residency status** |  |  |  |
| **Nursing home** | 2 (40) | 0 (0) | 6 (2) |
| **Non-institutional** | 3 (60) | 39 (98) | 324 (97) |
| **Unknown** | 0 (0) | 1 (3) | 4 (1) |
| **BMI (mean, SD)** | 33 (7) | 29.7 (6) | 29.37 (5.6) |
| **Comorbidity** |  |  |  |
| **Chronic pulmonary disease** | 1 (20) | 8 (20) | 91 (27) |
| **Chronic cardiac disease** | 2 (40) | 13 (33) | 71 (21) |
| **Immunocompromised** | 2 (40) | 8 (20) | 25 (7.5) |
| **Malignancy** | 0 (0) | 5 (13) | 27 (8) |
| **Obesity** | 3 (60) | 12 (30) | 128 (38) |
| **Diabetes** | 1 (20) | 18 (45) | 89 (27) |
| **Hypertension** | 2 (40) | 14 (35) | 110 (33) |
| **Other comorbidity** | 3 (60) | 24 (60) | 187 (56) |
| **Multi comorbidity** | 2 (40) | 21 (53) | 123 (37) |
| **No comorbidity** | 0 (0) | 1 (3) | 46 (14) |
| **Days since first vaccination, median (IQR)** | 60 [54, 70] | 29 [22, 36] | 5 [2, 8] |
| **Days since full vaccination, median (IQR)** | 33 [24, 35] | 4 [3, 6] | - |
| **Disease severity endpoints** |  |  |  |
| **Days in hospital (median [IQR])** | 6 [6, 7] | 6 [3, 9] | 6 [3, 10] |
| **ICU-admission (%)** | 0 (0) | 5 (13) | 64 (19) |
| **In-hospital mortality (%)** | 0 (0) | 9 (23) | 33 (10) |

## Discussion

We determined the effectiveness of COVID-19 vaccination against hospitalization, adjusting for comorbidity, during March 2021 to July 5 2021 in which the alpha variant was dominant in the Netherlands. The adjusted VE against hospitalization was 70% for partial vaccination and 93% for full vaccination. We found no significant differences between vaccine products, though the power for this analysis was low since 67% of the vaccinated patients received Comirnaty. We were not able to determine product specific estimates for COVID-19 Vaccine Janssen. There was protection against hospitalization for both full and partial vaccination in all comorbidity-subgroups. Only in immunocompromised patients the VE-point estimate was lower compared to the overall VE-estimate however, the confidence intervals were very wide. For all other subgroups, VE-point estimates for full vaccination were above 90%. Though VE estimates were generally higher in males, we found no significant effect of sex on VE. Analysis of secondary endpoints suggested less ICU-admission in fully vaccinated SARS-CoV-2 cases compared to partially vaccinated and unvaccinated cases, indicating additional protection of vaccination against ICU-admission compared to protection against hospitalization. However, this difference was not statistically significant due to the overall low number of patients needing ICU-admission.

VE estimates based on nationwide registration data in the Netherlands, where vaccination status among hospitalized COVID-19 cases is compared with vaccination coverage data from the population, yielded comparable VE estimates as in the current study. VE was 79% (95% CI: 77-80%) for partial vaccination and 94% (95% CI: 93-95%) for full vaccination during the period that the alpha variant was dominant (10). However, these estimates were not adjusted for sex and comorbidity. In addition, no distinction between hospitalization with or due to COVID-19 could be made based on the registration data, though the majority (an estimated 90%) was admitted due to COVID. Our findings are comparable to VE estimates against hospitalization during periods that the alpha variant was dominant reported from other countries, showing VE-estimates ranging from 33-74% for partial vaccination and 87-98% for full vaccination (11-14), and confirm the results of other studies also reporting no of large effects of adjustment for comorbidity on VE-estimates(11).

In one study performed in the United States, during the period of alpha-variant dominance, VE against hospitalization in immunocompromised patients was 63% (95% CI: 44-76%) as compared to 90% (95% CI: 87-92%) among subjects without immunocompromising conditions (14). Other studies have also reported lower immune response after vaccination among immunocompromised individuals (15-17). Among transplant patients the immune response was enhanced after a third vaccine dose (18, 19). Specific risk groups might, therefore, benefit from an additional vaccine dose. In the Netherlands, since the beginning of October a third vaccination is being offered to immunocompromised patients. Our study does not indicate other subgroups that might benefit from an additional vaccine dose in order to provide protection against COVID-19 requiring hospitalization, however, our study did not have enough power to detect small differences in VE for subgroups.

Whether waning will occur in the future or whether VE is affected by new variants such as the delta variant cannot be assessed by our study since the follow up time was too short. Therefore, follow up studies are necessary to guide policy making regarding booster vaccination.

Our findings are subject to a few limitations. First, we were not able to take into account immune status before hospitalization because serology was not consistently performed at admission, and prior SARS-CoV-2 infection was not consistently reported. This might have underestimated our VE-estimates. Second, our results are not representative for the VE against the delta variant, affecting the generalizability in the current epidemiologic situation. However, recent studies suggest that the VE against hospitalization is also very high in settings with dominance of the delta variant (10, 20).

Third, most patients were only partially vaccinated. Therefore, our study was underpowered to measure the effect of full vaccination in many subgroups. Fourth, it was not feasible to only include patients admitted because of suspected COVID-19 because SARS-CoV-2 PCR at admission is not reserved for patients suspected of a respiratory infection. Residual bias due to health-seeking behavior otherwise mitigated by the test negative design might therefore still exist. Based on the patient characteristics we suspect the control group to consist partially of chronically ill patients admitted for an acute deterioration of their respiratory or cardiac condition. This might have underestimated the VE since the sensitivity analysis showed higher VE estimates in a more infection-specific subgroup. Fifth, residual confounding due to unmeasured factors such as ethnicity might have biased our estimates. Sixth, there might be some misclassification of vaccination status by vaccinated patients who did not give informed consent for sharing their data with the national vaccination registry (7.3% of the Dutch population). We do not expect this effect to be large.

### Conclusion

We showed a high vaccine effectiveness of COVID-19 vaccination against COVID-19 related hospitalization during the period were de alpha variant was dominant. Also among risk groups, COVID-19 vaccination has a strong protective effect against hospitalization.

## Data Availability

Due to the safeguarding of the privacy of the subjects, case-based data is not publicly available

## Declarations

### Ethics approval

This study was performed in accordance with the ethical standards as laid down in the 1964 Declaration of Helsinki and its later amendments or comparable ethical standards. This study was assessed by the medical research ethics committee Utrecht and considered not to be subject to the Medical Research Involving Humans Subjects Act (WMO).

## Acknowledgements

The authors would like to acknowledge the hospitals that contributed to this study: Catharina Ziekenhuis, Medisch Spectrum Twente, Onze Lieve Vrouwe Gasthuis, Martini Ziekenhuis, St. Antonius Ziekenhuis and the Canisius Wilhelmina Ziekenhuis. In particular their research personnel that collected the data. Furthermore, the authors would like to acknowledge Jan van de Kassteele for his input on the statistical procedures.

---

**Table 1:**
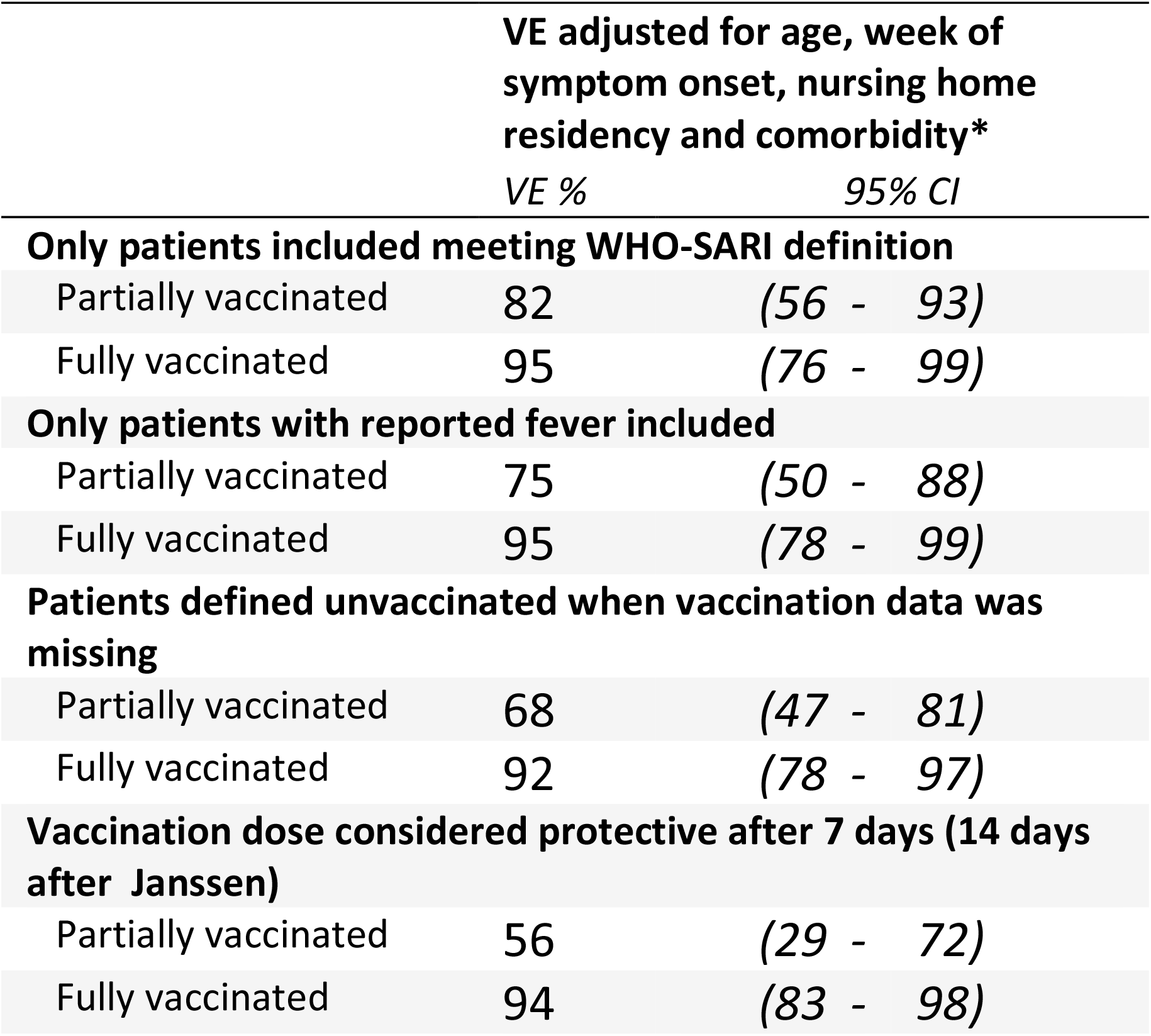
Sensitivity analysis

